# Oxytocin Enhances Social-Emotional Reciprocity in Autism

**DOI:** 10.1101/2025.07.20.25331870

**Authors:** Eric V. Strobl

## Abstract

We evaluated whether oxytocin improves social-emotional reciprocity in children and adolescents with autism spectrum disorder (ASD) by conducting a secondary, hypothesis-driven reanalysis of the SOARS-B trial, the largest randomized clinical trial of intranasal oxytocin to date involving over 272 youth. We used a machine learning approach to construct data-driven composite outcome measures maximally sensitive to treatment from the Aberrant Behavior Checklist modified Social Withdrawal (ABC-mSW) and Social Responsiveness Scale-2 Emotion Recognition (SRS2-ER) subscales. Permutation testing rigorously controlled the Type I error rate under outcome learning. Oxytocin significantly increased emotional responsiveness in the learned ABC-mSW composite (mean Cohen’s *d* = −0.25, *p* = 0.014) and improved social-emotional reciprocity in the learned SRS2-ER composite (mean *d* = −0.43, *p* = 0.022) compared to placebo. Both effects replicated in the open-label phase, with prior placebo participants also improving on the ABC-mSW composite (mean *d* = −0.30, *p* = 0.039) and the SRS2-ER composite (mean *d* = −0.22, *p* = 0.041) after crossing over to oxytocin. We thus provide robust evidence that oxytocin enhances social-emotional reciprocity in ASD – a rare finding given that most pharmacologic trials for core ASD symptoms are negative. The findings also highlight the value of outcome learning for detecting nuanced treatment effects and support oxytocin’s potential as a targeted intervention in ASD. Prospective, preregistered trials should replicate these effects to strengthen the evidence base.

**Lay Summary:** We found that the hormone oxytocin may help children and teens with autism improve how they share and respond to emotions with others. Using new computer methods, we discovered that oxytocin helped increase social connection and emotional responsiveness in ways that previous studies often missed. These results should be confirmed in new studies but suggest that oxytocin could be useful for improving social skills in autism.

## Introduction

Oxytocin is a neuropeptide critical for social cognition and affiliation across species, with robust preclinical evidence demonstrating its role in social recognition, bonding, and approach behaviors (Pedersen and Prange Jr, 1979; Young and Wang, 2004; Ferguson et al., 2001; Kosfeld et al., 2005; Domes et al., 2007; Kirsch et al., 2005). As a result, oxytocin emerged as a promising candidate for addressing the core social deficits of autism spectrum disorder (ASD). Observational studies strengthened this rationale by identifying altered endogenous oxytocin levels and oxytocin receptor gene variants in individuals with ASD, which helped motivate the launch of multiple randomized clinical trials (RCTs) (Modahl et al., 1998; Wu et al., 2005). However, results from early RCTs were mixed: some found small, inconsistent improvements in social cognitive domains such as emotion recognition, social gaze, and reciprocity, but most effects were modest and failed to replicate (Guastella et al., 2010; Anagnostou et al., 2012). Other studies reported no significant benefit over placebo on primary ASD symptoms or overall social functioning (Dadds et al., 2014; Watanabe et al., 2015). As a result, oxytocin’s efficacy for ASD social deficits remained uncertain, with reported effects generally small, inconsistent, and lacking robust replication.

The SOARS-B trial was a multi-site, placebo-controlled, phase II study that addressed the uncertainty by evaluating 24 weeks of intranasal oxytocin in 277 children and adolescents with ASD after exclusions (Sikich et al., 2021). The primary outcome was the Aberrant Behavior Checklist modified Social Withdrawal (ABC-mSW) subscale (Aman et al., 1985; Sikich et al., 2021), supplemented by secondary measures targeting social motivation and broader social functioning, such as the Social Responsiveness Scale-2 (SRS2) (Constantino, 2013). Unfortunately, the oxytocin and placebo groups exhibited nearly identical changes on all primary and secondary endpoints, with no statistically significant evidence of benefit from oxytocin in any analyzed subgroup, including those stratified by age or verbal fluency. Sensitivity analyses did not alter the results. Thus, despite robust preclinical rationale and prior positive pilot data, the SOARS-B trial found no evidence that longer-term intranasal oxytocin improves core social functioning in ASD.

Social functioning in ASD is, however, a multidimensional construct, encompassing domains such as emotion recognition, social avoidance, and interpersonal relatedness (Frazier et al., 2014). Total scores and even sub-scores of instruments like the ABC and SRS2 are relatively coarse and thus may not capture more nuanced or domain-specific effects of interventions like oxytocin. In particular, if oxytocin’s effects are specific to social-emotional reciprocity without broadly improving most other aspects of social functioning, then traditional fixed outcome measures can obscure real but subtle treatment effects. To address this challenge, we re-analyzed the SOARS-B dataset using a recently developed machine learning approach that identifies composite measures most sensitive to treatment effects, enabling the detection of nuanced, domain-specific improvements that conventional analyses may miss (Strobl, 2025). We focused on social-emotional reciprocity because oxytocin’s established role in strengthening the maternal-infant bond depends critically on this domain (Feldman et al., 2007; Feldman, 2012).

We hypothesized that oxytocin would selectively enhance social-emotional reciprocity in ASD, leading to reductions in maladaptive behaviors specifically linked to limited social engagement, as captured by a learned, data-driven ABC-mSW composite. We further posited that increased reciprocity would diminish deficits and facilitate gains in adaptive social functioning, as reflected in improved performance on a learned composite derived from the SRS2 Emotion Recognition (SRS2-ER) subscale (Frazier et al., 2014), which includes items targeting emotional responsiveness and social flexibility. Importantly, we expected these improvements to accrue gradually over the course of 12 weeks, as participants developed new skills in response to enhanced social-emotional reciprocity.

## Methods

### SOARS-B Trial Design

We performed a secondary analysis of the Study of Oxytocin in Autism to improve Reciprocal Social Behaviors (SOARS-B) trial (ClinicalTrials.gov ID NCT01944046; (Sikich et al., 2021)). The SOARS-B trial was a multicenter, randomized, double-blind, placebo-controlled, phase II study evaluating the efficacy and safety of daily intranasal oxytocin in children and adolescents with ASD. Participants were randomized to receive either oxytocin or placebo for 24 weeks, with outcomes assessed using standardized measures of social functioning and adaptive behavior, such as the ABC and SRS2. Oxytocin was administered intranasally to a target dose of 48 international units (IU) per day dosed 24 IU twice daily. The target dose could be increased by 16 IU increments every 4 weeks, up to a maximum of 80 IU per day, after maintaining the initial target dose for 7 weeks. Following the double-blind phase, all participants proceeded to a 24-week open-label extension in which they received active oxytocin with knowledge of their treatment assignment. The ABC was administered every 4 weeks across both phases (weeks 0-48), while the SRS2 was assessed every 12 weeks.

We downloaded the SOARS-B dataset from the [National Institute of Mental Health Data Archive](https://nda.nih.gov/) with a limited access data use certificate awarded to author Eric V. Strobl.

### Eligibility Criteria

Trial investigators enrolled children and adolescents aged 3 to 17 years who met DSM-5 criteria for ASD, as confirmed through clinical interviews, neurological and physical examinations, cognitive assessments (Stanford-Binet Intelligence Scales, Fifth Edition (Roid and Pomplun, 2012), or Mullen Scales of Early Learning (Mullen et al., 1995)), and administration of the Autism Diagnostic Observation Schedule, Second Edition (ADOS-2) (Lord et al., 2012). Investigators required parents or guardians to speak English. They excluded individuals with Rett syndrome, childhood disintegrative disorder, deafness, blindness, active cardiovascular or renal disease, uncontrolled epilepsy, pregnancy, lactation, or sexual activity without contraception. Investigators also excluded individuals with a history of daily intranasal oxytocin use for more than 30 days. They required neuropsychiatric medication regimens to remain stable for at least one month before randomization, and nonmedication ASD therapies to remain unchanged for at least two months. Trial investigators permitted the use of antipsychotic agents, anticonvulsants, and stimulants during the trial.

### Outcome Learning

Standard analytic approaches in ASD trials typically employ fixed scores from established instruments such as the ABC or SRS2. However, both total and subscale scores may lack sensitivity to intervention effects that are restricted to specific dimensions or subdomains of social functioning. Given the multidimensional nature of sociality in ASD, interventions like oxytocin may produce subtle or domain-specific effects that conventional outcome measures dilute or overlook (Brugha et al., 2015; McConachie et al., 2015). Exclusive reliance on fixed composite scores can therefore obscure genuine but targeted treatment benefits, particularly when overall scales include items unrelated to the intervention’s expected mechanism of action. These limitations are now widely recognized; the Lancet Commission on Autism explicitly called for outcome measures that are sensitive to domain-specific change, highlighting the inadequacy of current tools for capturing real-world improvement in autism intervention trials (Lord et al., 2022).

We adopted a composite outcome learning approach to address the above limitations. Specifically, we used the recently developed Sparse Canonical Outcome Regression (SCORE) algorithm (Strobl, 2025) to construct data-driven composite outcomes that are maximally sensitive to oxytocin’s effects relative to placebo. SCORE identifies a sparse, non-negative weighting of individual items from a clinical rating scale (e.g., the ABC-mSW or SRS2-ER), resulting in a targeted composite that best distinguishes treatment groups as quantified by Cohen’s *d*. This strategy enhances both interpretability and power to detect clinically meaningful changes that might otherwise be obscured by noise from unrelated items. We thus conducted all analyses with SCORE, assuming missingness at random (MAR) (Rubin, 1976) and using all available data points with complete item-level data at baseline and at least one follow-up time point. We did not impute any data.

### Endpoints and Nuisance Variables

We defined the primary endpoint as Cohen’s *d* averaged over weeks 12, 16, 20, and 24 in the 13 items comprising the ABC-mSW, a caregiver-rated measure of maladaptive behaviors related to social withdrawal and reduced social engagement(Sikich et al., 2021; Aman et al., 1985). The ABC-mSW was the prespecified primary outcome measure in SOARS-B. This 12-week evaluation window provides sufficient time for oxytocin’s effects to emerge, and averaging across multiple late-phase assessments maximizes statistical power while minimizing the influence of measurement variability. We hypothesized that oxytocin would reduce maladaptive behaviors related to decreased social-emotional reciprocity by week 12 and sustain this therapeutic effect throughout the remaining 12 weeks.

If oxytocin exerts meaningful effects, we should observe not only a reduction in maladaptive behaviors but also an increase in adaptive behaviors related to social-emotional reciprocity. We thus assessed both maladaptive and adaptive social behaviors using the SRS2-ER subscale as a secondary measure. This subscale was introduced by Frazier et al. (2014) as part of an empirically derived five-factor structure. The five-factor solution showed superior fit to one-to four-factor and hierarchical alternatives in a large confirmatory factor analysis of 9,635 children and adults with and without ASD. Moreover, the five-factor structure demonstrated (1) *known-groups validity* with consistent separation between ASD and unaffected siblings, (2) *genetic coherence* with larger monozygotic twin-twin correlations than dizygotic correlations, (3) *discriminant structure* with non-uniform inter-factor correlations, and (4) *construct validity* with exclusive mappings to one of the two DSM-5 ASD symptom domains. Within this structure, ER captures the capacity to recognize, interpret, and appropriately react to social-emotional cues from others, making it a suitable, psychometrically supported endpoint for evaluating oxytocin’s effects on social-emotional reciprocity.

The original investigators classified patients as minimally verbal if they completed ADOS-2 modules 1 or 2, and as fluently verbal if they completed modules 3 or 4 (Sikich et al., 2021). All analyses included age, sex, and verbal fluency status as nuisance variables. We inherently adjusted for baseline scores by analyzing the change from baseline in each item of the ABC-mSW and SRS2-ER, ensuring that all effects were estimated relative to individual starting points.

### Open-Label Replication

The SOARS-B trial offers a unique opportunity for within-trial replication through its 24-week open-label extension, during which all participants received oxytocin. We derived the composite outcomes exclusively from the randomized phase data and then applied them, without modification, to the open-label extension. If oxytocin is effective, we would expect participants originally assigned to placebo to exhibit declines in maladaptive behaviors – as measured by the ABC-mSW composite learned in the randomized phase – after crossing over to oxytocin. Similarly, improvements should emerge in both maladaptive and adaptive social behaviors on the SRS2-ER composite in this group. We therefore hypothesized that, beginning at week 36 (12 weeks into the open-label phase), the former placebo group would demonstrate not only reduced maladaptive behaviors but also gains in adaptive skills indicative of enhanced social-emotional reciprocity relative to their status at the end of the randomized phase (week 24).

### Hypothesis Testing

We used permutation testing to ensure valid hypothesis testing after estimating the outcome weights. We assumed that treatment assignments (oxytocin or placebo) were exchangeable under the null hypothesis. We permuted treatment labels 10,000 times to approximate the null distribution of our test statistic, calculating mean Cohen’s *d* across weeks 12 to 24 for the learned composite outcome in each permutation. We then compared the observed mean Cohen’s *d* to this null distribution, and defined the p-value as the proportion of permutations with a test statistic as extreme or more extreme than the observed value. We did not adjust for multiple hypothesis testing, as we pre-specified a single composite outcome per endpoint.

For the open-label phase, we evaluated whether patients originally assigned to placebo showed improvement between weeks 24 and 36-48 after crossing over to oxytocin. We assumed that week 24 and weeks 36-48 were exchangeable under the null hypothesis. We thus randomly permuted the week 24 and the weeks 36-48 block within subjects 10,000 times and recalculated mean Cohen’s *d* across weeks 36-48 for each permutation. We again computed the p-value as the proportion of permutations yielding a test statistic as or more extreme than the observed value.

In Supplementary Materials, we provide empirical evidence that pairing SCORE with the permutation testing procedures described above does not generate false positives in this trial or in an independent trial, thereby demonstrating proper control of the Type I error rate despite outcome learning.

### Statistical Software

All statistical analyses were performed in R version 4.3.1. The SCORE algorithm was implemented in R, with source code available at github.com/ericstrobl/SCORE.

### Use of Artificial Intelligence Tools

We did not use artificial intelligence or large language model tools to generate content for this manuscript.

## Results

### Participant Characteristics

The original SOARS-B trial recruited 290 patients. Guardians for six patients declined permission for their child’s data to be uploaded to the NIMH Data Archive. Of the remaining 284 patients, two lacked baseline data and ten lacked data at all follow-up time points. Consequently, our analysis included 290 − 6 − 2 − 10 = 272 patients. The shared dataset lacked racial and ethnic identifiers, as well as plasma oxytocin measurements, for all participants. Table 1 summarizes the available characteristics at baseline.

**Table 1:** Patient characteristics of the SOARS-B dataset. We summarize continuous variables as mean (standard deviation) over all non-missing data. We present count (percentage) for biological sex. 1The ABC Social Withdrawal subscale is also known as the Lethargy subscale. 2SRS2 subscales were derived from the confirmatory factor analysis results reported in (Frazier et al., 2014). To ensure consistency in interpretation, adaptive items were recoded as maladaptive by inverting their scores – multiplying by negative one and adding the maximum possible score – so that higher values consistently reflect greater symptom severity.

|  | All Participants |  | Minimally Verbal |  | Fluently Verbal |  |
| --- | --- | --- | --- | --- | --- | --- |
|  | Oxytocin<br>(*n*=138) | Placebo<br>(*n*=134) | Oxytocin<br>(*n*=65) | Placebo<br>(*n*=65) | Oxytocin<br>(*n*=73) | Placebo<br>(*n*=69) |
| Demographics |  |  |  |  |  |  |
| Age | 10.4 (4.05) | 10.4 (3.98) | 9.75 (3.96) | 9.78 (3.66) | 11.1 (4.05) | 11.1 (4.18) |
| Sex (male) | 121 (87.7) | 116 (86.6) | 56 (86.2) | 54 (83.1) | 65 (89.0) | 62 (89.9) |
| ABC Subscales |  |  |  |  |  |  |
| mSW | 11.0 (6.91) | 11.5 (7.63) | 11.9 (7.06) | 13.4 (7.50) | 10.2 (6.72) | 9.81 (7.40) |
| Irritability | 10.4 (7.40) | 11.7 (8.36) | 10.5 (6.97) | 11.9 (8.58) | 10.2 (7.82) | 11.6 (8.20) |
| Social Withdrawal<sup>1</sup> | 11.8 (7.61) | 12.7 (8.43) | 12.5 (7.47) | 14.6 (8.20) | 11.3 (7.74) | 10.9 (8.29) |
| Stereotypy | 5.87 (4.90) | 5.67 (4.83) | 7.49 (5.41) | 7.18 (5.12) | 4.40 (3.86) | 4.25 (4.09) |
| Hyperactivity | 19.4 (11.0) | 19.1 (10.3) | 20.4 (11.8) | 21.5 (10.8) | 18.6 (10.3) | 16.8 (9.39) |
| Inappropriate Speech | 4.22 (3.23) | 4.57 (3.27) | 3.95 (3.48) | 5.11 (3.51) | 4.47 (3.00) | 4.06 (2.96) |
| SRS2 Subscales<sup>2</sup> |  |  |  |  |  |  |
| Emotion Recognition | 53.7 (10.3) | 54.2 (9.46) | 57.0 (10.4) | 58.0 (7.99) | 50.7 (9.34) | 50.6 (9.39) |
| Social Avoidance | 14.4 (4.25) | 14.9 (4.25) | 14.3 (4.24) | 15.9 (4.38) | 14.4 (4.29) | 14.0 (3.94) |
| Interpersonal Relatedness | 16.0 (3.57) | 16.0 (3.66) | 15.6 (3.90) | 16.2 (3.89) | 16.4 (3.21) | 15.9 (3.45) |
| Insistence on Sameness | 44.6 (8.47) | 45.1 (8.77) | 42.5 (8.91) | 44.7 (9.14) | 46.4 (7.66) | 45.4 (8.45) |
| Repetitive Mannerisms | 20.3 (5.13) | 20.1 (5.27) | 22.1 (5.45) | 22.4 (5.29) | 18.7 (4.25) | 17.8 (4.20) |
| Stanford Binet 5 |  |  |  |  |  |  |
| Abbreviated IQ | 85.1 (23.6) | 82.0 (22.9) | 66.8 (18.8) | 61.7 (13.7) | 94.5 (20.0) | 93.2 (19.0) |

The average age of participants was approximately 10 years, and about 87% were male. At baseline, minimally verbal patients exhibited a higher frequency of maladaptive behaviors related to social withdrawal, as measured by the ABC-mSW subscale. Minimally verbal patients also displayed more pronounced deficits in emotion recognition, as assessed by the SRS2-ER. We therefore included verbal fluency status, age, and sex as covariates in all subsequent analyses.

### Primary Endpoint

We first replicated prior results, confirming that the standard ABC-mSW subscore did not distinguish oxytocin from placebo (mean Cohen’s *d* = −0.01, *p* = 0.432; Figure 1 (a)); we plot all raw scores in Supplementary Figure 1. However, oxytocin may affect specific aspects of social withdrawal not captured by the overall score. To test this, we constructed a data-driven weighted composite from all 13 ABC-mSW items. This approach identified two items – lack of emotional responsiveness and showing few social reactions to others – as most sensitive to treatment. The learned item weights are shown in Figure 1 (b). Using this composite, oxytocin demonstrated a significantly greater reduction in maladaptive behaviors than placebo from weeks 12 to 24 (mean Cohen’s *d* = −0.25, *p* = 0.014; Figure 1 (c)). These results indicate that oxytocin specifically reduced behaviors reflecting diminished emotional expressivity, highlighting the value of targeted, item-level outcome analysis.

**Figure 1.**
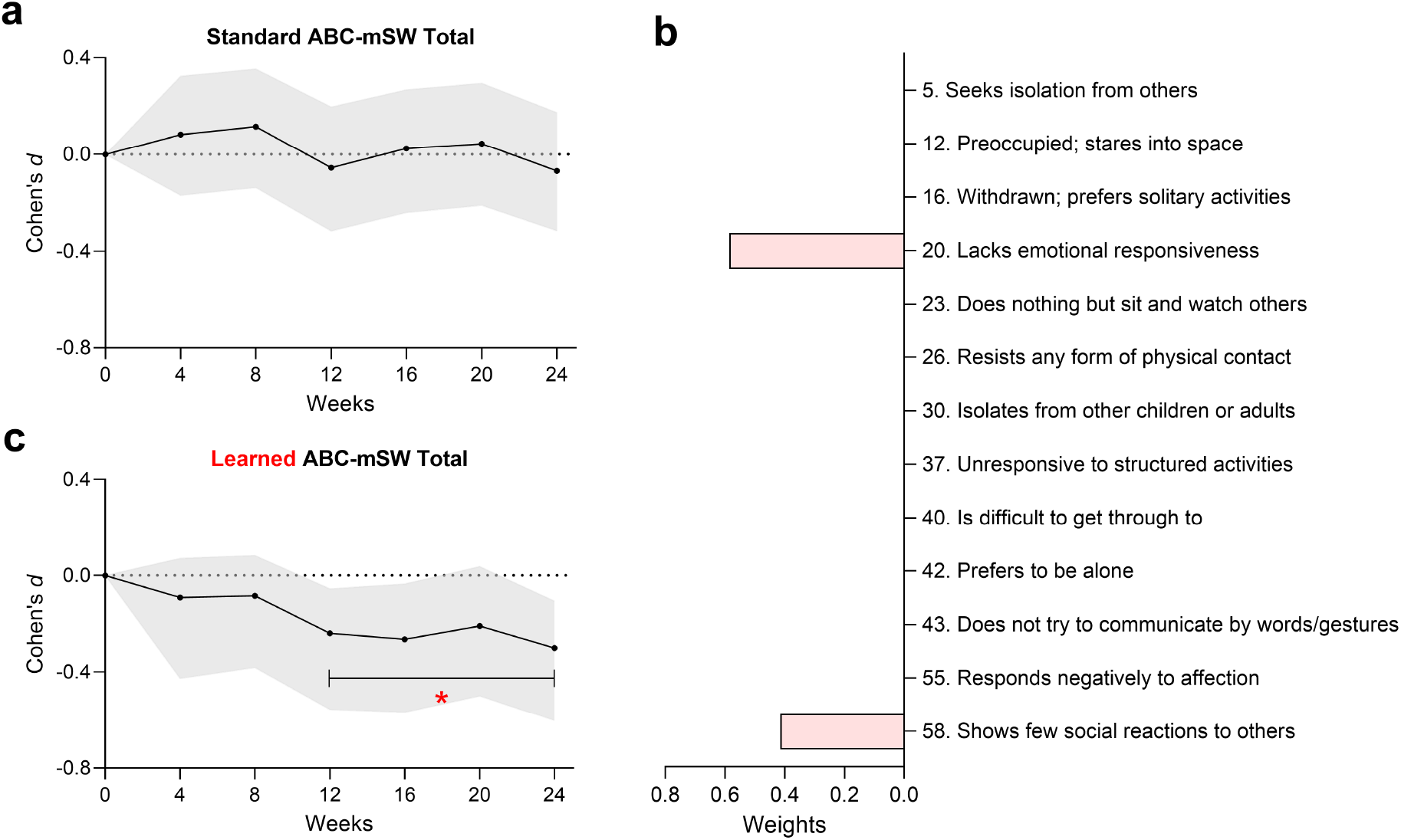
Effects on maladaptive behaviors as assessed by the ABC-mSW. (a) We successfully replicated prior findings, observing no significant differences between treatment groups at any measured time point using the total ABC-mSW score. (b) Given that the ABC-mSW comprises 13 items, we constructed an optimally weighted composite score that maximally distinguished oxytocin from placebo, as quantified by Cohen’s *d*. This data-driven weighting prioritized only two closely related items: lack of emotional responsiveness and showing few social reactions to others. (c) Oxytocin significantly reduced the weighted combination of these two items from weeks 12 to 24. The horizontal interval bars do not represent a comparison between the endpoints; instead, they indicate that the mean value across the time points within each interval differs significantly in the oxytocin group compared to placebo. Unless indicated otherwise, shaded error bands represent 95% confidence intervals in this paper, estimated at each time point using 10,000 bootstrap resamples. * *p <* 0.05, ** *p <* 0.01, *** *p <* 0.001 based on permutation testing with 10,000 permutations. Both the bootstrap and permutation procedures accounted for the adaptive selection of weights.

### Secondary Endpoint

While the ABC-mSW assesses maladaptive behaviors linked to social withdrawal, improvements in emotional expressivity may also manifest as gains in both maladaptive and adaptive domains, as measured by the SRS2-ER subscale. We first confirmed that the standard SRS2-ER subscale did not distinguish oxytocin from placebo with equal weighting of all 25 items (mean Cohen’s *d* = −0.06, *p* = 0.260; Figure 2 (a)). We then learned a composite outcome from the emotion recognition items that best differentiated treatment groups. The SCORE algorithm identified 6 items that were most sensitive to oxytocin’s effects, each reflecting active social-emotional reciprocity (Figure 2 (b)). This targeted composite revealed a significant and sustained treatment effect at 12 and 24 weeks (mean Cohen’s *d* = −0.43, *p* = 0.022; Figure 2 (c)), indicating that oxytocin decreased maladaptive and enhanced adaptive behaviors related to social-emotional reciprocity.

**Figure 2.**
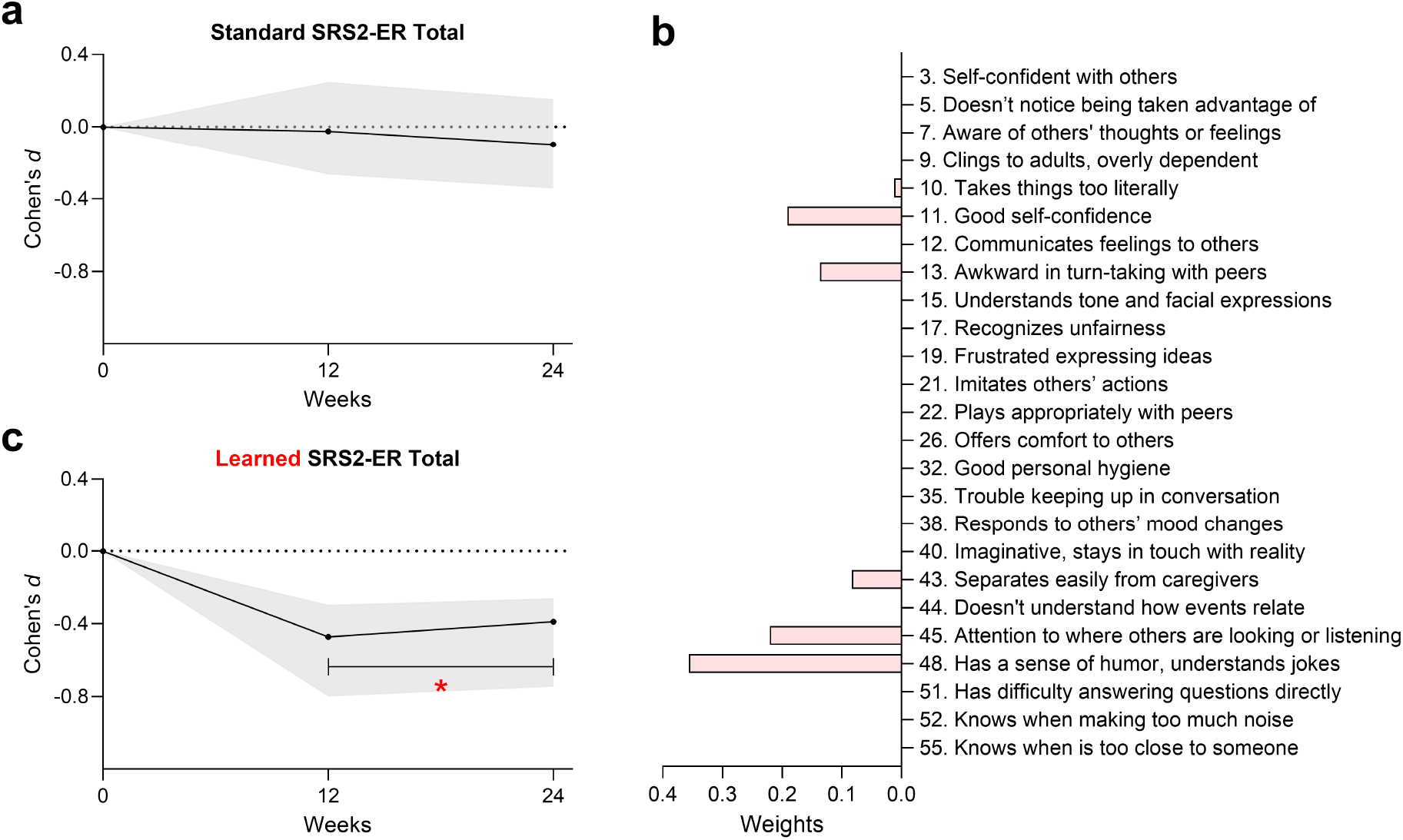
Effects on maladaptive and adaptive behaviors as assessed by the SRS2-ER. (a) Equal weighting of all SRS2-ER items failed to distinguish oxytocin from placebo. (b) Outcome learning identified a composite outcome comprising a weighted sum of only 6 out of the 25 SRS2-ER items. (c) Oxytocin produced a significant therapeutic effect on this composite at weeks 12 and 24, mirroring the results observed with the ABC-mSW composite.

### Open-Label Replications

We next analyzed the open-label phase of the SOARS-B trial, in which all participants knowingly received oxytocin. If oxytocin exerts a true therapeutic effect, then applying the ABC-mSW composite – learned solely from the randomized phase – should reveal declines in maladaptive behaviors among participants who switched from placebo to oxytocin in the open-label phase. Consistent with this hypothesis, these patients exhibited a significant decrease in maladaptive behaviors during the open-label phase relative to the end of the randomized phase (mean Cohen’s *d* = −0.30, *p* = 0.039; Figure 3 (a)). We likewise anticipated decreased maladaptive and enhanced adaptive behaviors as measured by the SRS2-ER composite. Placebo patients indeed demonstrated a significant decrease in the SRS2-ER composite following open-label oxytocin (mean Cohen’s *d* = −0.22, *p* = 0.041; Figure 3 (b)). Note that we also observed significant improvements in the original total scores, but these did not replicate in the randomized phase (Supplementary Figure 2). Finally, visual inspection of Figures 1 (c) and 2 (c) indicated that oxytocin’s effect plateaued by week 12. Consistent with this, patients assigned to oxytocin showed no additional improvement between week 24 and weeks 36-48 during the open-label phase: ABC-mSW (mean Cohen’s *d* = −0.11, *p* = 0.164) and SRS2-ER (mean Cohen’s *d* = 0.04, *p* = 0.435).

**Figure 3.**
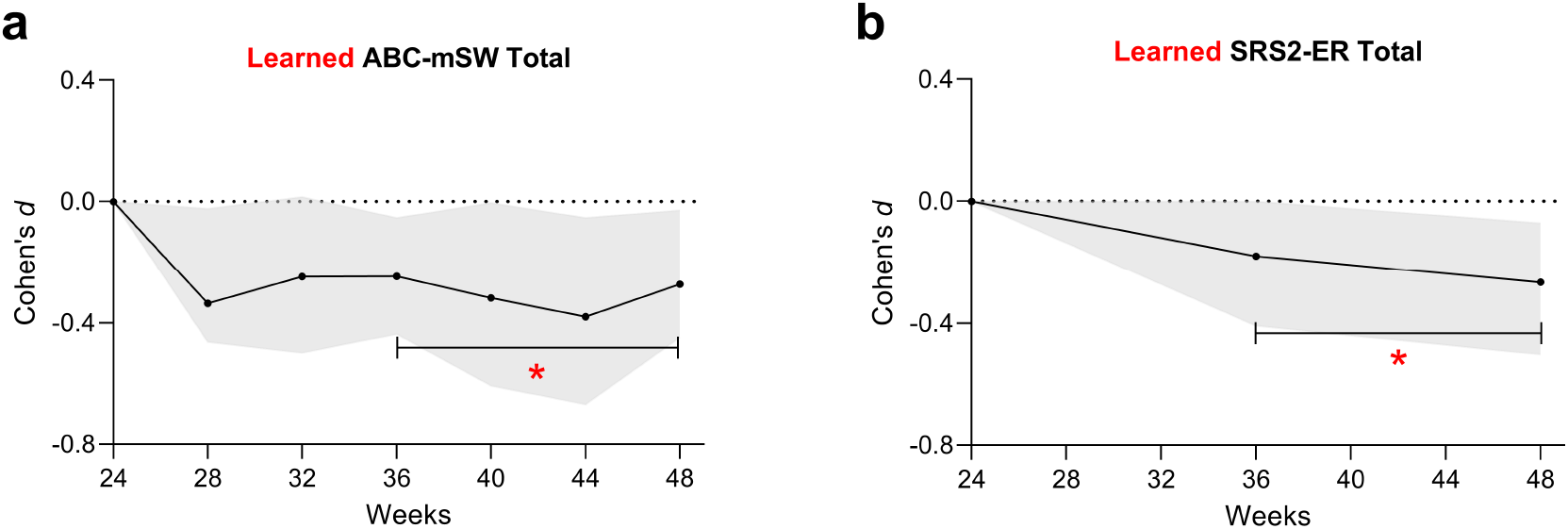
Effects in the open-label extension phase. (a) Oxytocin significantly reduced maladaptive behaviors, as measured by the ABC-mSW composite, at weeks 36-48. (b) Oxytocin also produced significant improvements in both maladaptive and adaptive behaviors based on the SRS2-ER composite.

## Discussion

We re-analyzed data from the SOARS-B trial to assess the effects of oxytocin on social-emotional reciprocity in ASD, applying machine learning to derive optimized composite outcome measures. Oxytocin led to significant reductions in maladaptive behaviors associated with diminished emotional expressivity and social reactivity, observed both in the oxytocin group during the initial 24-week randomized phase and in the former placebo group during the open-label extension. Notably, oxytocin’s benefits extended beyond mitigating maladaptive behaviors; we also observed sustained improvements in adaptive social behaviors, such as humor, attention to others, self-confidence, and turn-taking. These adaptive gains were also observed in the placebo group after crossover to open-label oxytocin. Our results thus indicate that oxytocin not only reduces maladaptive behaviors but also enhances adaptive behaviors related to social-emotional reciprocity in ASD.

The observed improvements in social-emotional reciprocity are biologically plausible given oxytocin’s established role in modulating social behavior and affiliative processes. Oxytocin receptors are densely localized in the human amygdala and anterior cingulate, with evidence for similar distributions in limbic and hypothalamic regions (Boccia et al., 2013). Intranasal oxytocin administration has been shown in pharmacological fMRI studies to acutely attenuate amygdala reactivity to social stimuli, especially fear-inducing facial expressions, and to alter functional connectivity between the amygdala and brainstem regions implicated in fear responses (Kirsch et al., 2005). These neural effects are hypothesized to underlie oxytocin’s capacity to reduce social threat perception and promote trust. Beyond reducing anxiety, oxytocin also increases the salience of social cues and promotes attention to the eye region of faces in humans, while animal and translational studies support additional roles in facilitating the encoding of positive social memories and modulating reward-related neural circuits (Meyer-Lindenberg et al., 2011). This convergence of anxiolytic, salience-enhancing, and affiliative effects provides a mechanistic foundation for oxytocin’s capacity to both diminish maladaptive social withdrawal and foster adaptive social engagement, as observed in our study.

Clinically, enhancing social-emotional reciprocity is significant for several reasons. Increases in this domain can strengthen caregiver-child connections and potentially reduce caregiver burnout – a critical concern given the long-term demands of caring for individuals with ASD (Dückert et al., 2023). Improved social-emotional reciprocity also enables children to share emotions, respond to social cues, and participate in reciprocal interactions, fostering more meaningful relationships and engagement with peers and caregivers (Yizengaw, 2022). Such gains can reduce social isolation, support language and adaptive skill development, and enhance participation in educational and community settings. Notably, increases in social-emotional reciprocity are associated with reduced risk of comorbid psychiatric symptoms in ASD – particularly social anxiety and behavioral challenges – as greater improvements in social responsiveness predict significant reductions in anxiety symptoms, and deficits in social reciprocity are correlated with heightened anxiety and behavioral difficulties (Factor et al., 2022; Fuselier et al., 2024). Finally, longitudinal studies indicate that early improvements in reciprocity are among the strongest predictors of better adaptive functioning and community integration in adulthood (Howlin et al., 2013).

Our findings were made possible by leveraging outcome learning (Strobl, 2025), which empirically identified composite scores most sensitive to oxytocin’s effects, rather than relying exclusively on fixed outcomes. Conventional measures, such as the ABC-mSW and SRS2-ER subscores, did not yield significant group differences in our study and in many past studies because they dilute true intervention effects by assigning equal weight to all items (Sikich et al., 2008; Guastella et al., 2023). We instead detected subtle yet clinically relevant treatment effects by applying data-driven weights to individual items; rigorous permutation testing at each stage ensured valid inference and appropriate control of the Type I error rate, despite the outcome learning process. Our results ultimately demonstrate that machine learning can substantially refine outcome measurement by starting with broad subscores and optimizing them based on empirical data, thereby uncovering subtle but significant treatment effects that standard scores may overlook.

The open-label findings further corroborated the randomized-phase results because a genuine treatment effect should manifest in both the randomized and open-label settings. The learned ABC-mSW and SRS2-ER composites exhibited significant effects in both phases, demonstrating sensitivity to causal effects under randomization and to treatment-related change under nonblinded, real-world conditions. By contrast, the original total scores reached significance only in the open-label extension, consistent with improvement driven primarily by open-label biases such as expectancy and ascertainment. Moreover, open-label improvement is typically easier to detect on broad total scores than on sparse, few-item composites. Total scores aggregate nonspecific shifts and are more vulnerable to the *halo effect*, in which perceived improvement in one item inflates ratings elsewhere(Dayan et al., 2024). Sparse composites are less susceptible because they rely on fewer items, so observing open-label improvement on a sparse composite constitutes a stricter test of meaningful change. Accordingly, we found the strongest support for oxytocin’s effectiveness in SOARS-B by observing significant effects on a sparse composite in *both* the randomized and open-label phases.

Several limitations warrant mention. First, as this is a secondary analysis, the findings should be interpreted with caution and confirmed in independent prospective studies to guard against chance discoveries. However, we rigorously applied permutation testing at each analytic stage and internally replicated the primary outcome results within the SOARS-B trial across three analyses: in the SRS2-ER secondary endpoint during the randomized phase, and in both the ABC-mSW and SRS2-ER composites during the open-label phase. Second, we did not stratify our analyses by patient subgroups – such as genetic profiles or baseline characteristics – because our primary aim was to establish foundational insights into the effects of oxytocin across a diverse cohort with ASD. Stratified analyses may indeed reveal larger effects within specific populations, but such analyses depend on first establishing at least some overall effects in the full sample, which provides the necessary foundation for meaningful subgroup discovery in future work. Third, our analyses relied on standard rating scales such as the ABC and SRS2, which may not fully capture the nuanced and context-dependent nature of social reciprocity in ASD, even with outcome learning. As a result, the subscales may potentially underestimate the domain-specific effects of oxytocin. Fourth, the observed effect sizes for oxytocin were small to moderate, with mean Cohen’s *d* values of −0.25 for the ABC-mSW composite and −0.43 for the SRS2-ER composite during the randomized phase. Consequently, it remains uncertain whether these statistical improvements correspond to meaningful changes in daily social functioning, interpersonal relationships, or overall quality of life – outcomes of primary importance in psychiatric care and for families. Finally, we cannot determine whether improvements persist after discontinuing oxytocin, as follow-up beyond the treatment period was not conducted. Future work should thus prioritize independent replication, patient stratification to identify subgroups most likely to benefit, the development of more sensitive and ecologically valid outcome measures, and longer-term follow-up to determine the durability and real-world significance of treatment effects.

In summary, our study provides evidence that oxytocin improves social-emotional reciprocity in ASD when assessed using a data-driven outcome learning approach. These findings suggest that oxytocin targets highly specific facets of social behavior, supporting its potential use as a targeted intervention in ASD.

## Data Availability

We downloaded the SOARS-B dataset from the National Institute of Mental Health Data Archive with a limited-access data use certificate.

## Acknowledgments

None

## Supplementary Materials

### Additional Verification

We evaluated SCORE with permutation tests in scenarios where one treatment was believed to be inferior to or equivalent with another on *every* item of the relevant symptom rating scale. To guarantee item-wise inferiority or equivalence, we restricted attention to treatment pairs where one could be viewed as a subset of the other without any opposing effects on any item. For instance, we required outcome learning to search for item sets that would spuriously make medication appear superior to medication plus therapy, or placebo superior to medication. We excluded contrasts such as a non-sedating medication versus a non-sedating plus a sedating medication, because the non-sedating medication alone may be more effective for hypersomnia, producing a genuine item-level advantage. Under these constraints, a valid procedure should not discover a more beneficial effect for the inferior or equivalent treatment, thereby demonstrating control of the Type I error rate on real data.

We first forced SCORE to construct outcomes that show superiority of placebo over oxytocin in the SOARS-B trial. After swapping oxytocin and placebo labels and repeating the permutation test, we observed no differences between oxytocin and placebo for ABC-mSW (mean Cohen’s *d* = −0.18, *p* = 0.130) and SRS2-ER (mean Cohen’s *d* = −0.25, *p* = 0.541). Nonsignificant results also held in the open-label phase: ABC-mSW (mean Cohen’s *d* = −0.09, *p* = 0.448) and SRS2-ER (mean Cohen’s *d* = 0.01, *p* = 0.708). These analyses indicate that the permutation procedure did not spuriously favor placebo over oxytocin.

We next assessed performance in an independent randomized clinical trial called the Treatment for Adolescents with Depression Study (TADS). The trial compared three treatment arms – fluoxetine, cognitive-behavioral therapy (CBT), and combination therapy – for 36 weeks using the Children’s Depression Rating Scale-Revised (CDRS-R)(March et al., 2007). We instructed SCORE to spuriously learn outcomes favoring each solo treatment over combination therapy at weeks 24-36 because we do not expect fluoxetine or CBT alone to outperform combination therapy. As expected, the procedure did not find any solo arm superior to combination: fluoxetine (mean Cohen’s *d* = −0.14, *p* = 0.601) and CBT alone (mean Cohen’s *d* = −0.27, *p* = 0.132). Conversely, the algorithm readily recovered the superiority of combination therapy over both fluoxetine (mean Cohen’s *d* = −0.41, *p* = 0.004) and CBT alone (mean Cohen’s *d* = −0.44, *p* = 0.003), even though the original trial reported no significant difference between fluoxetine and combination therapy – a null finding we replicated (mean Cohen’s *d* = −0.14, *p* = 0.086). These results highlight the enhanced statistical power of outcome learning and provide further support that the permutation framework controls the Type I error rate despite outcome learning.

### Raw Scores

**Supplementary Fig. 1:**
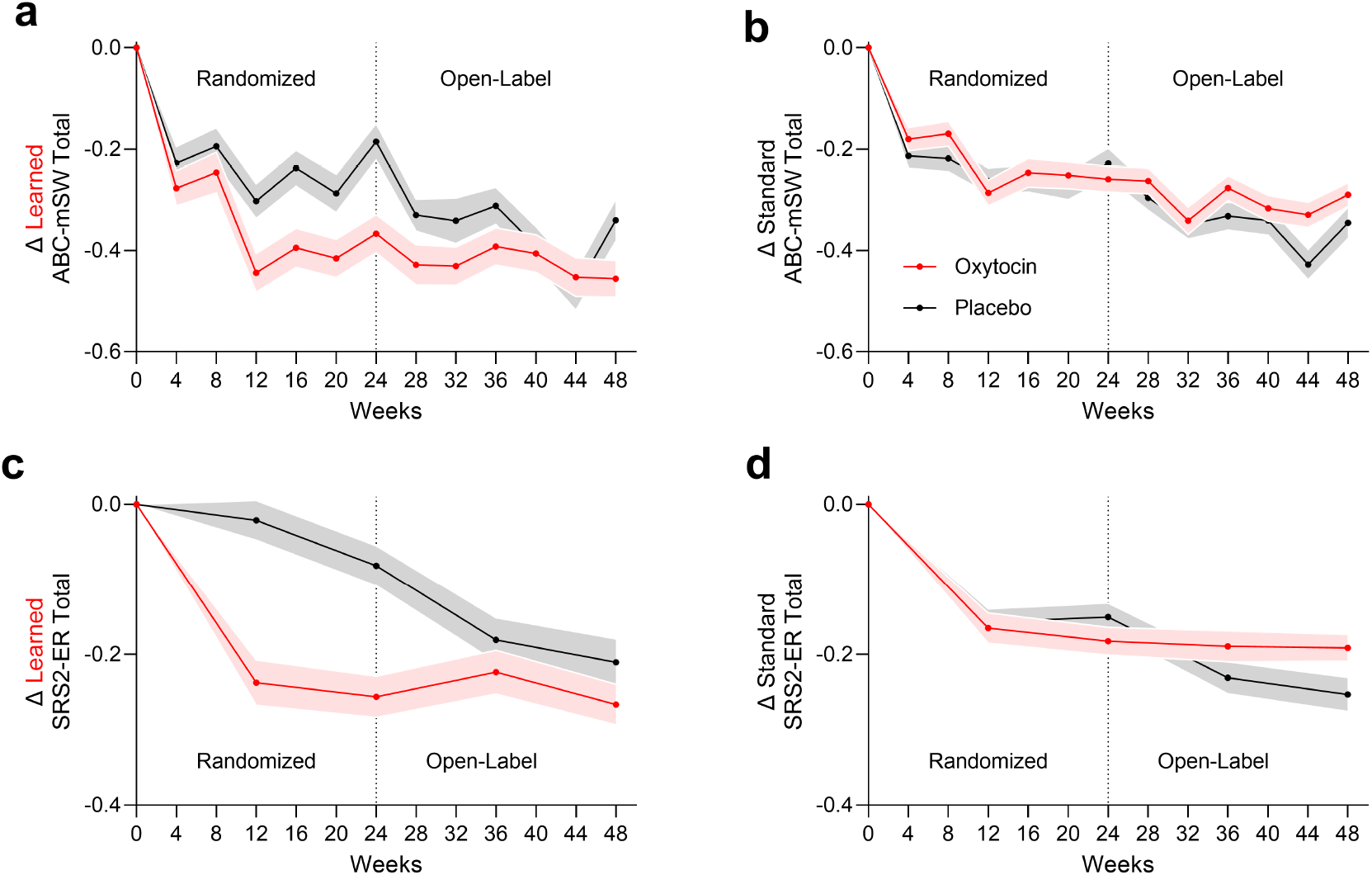
Raw change from baseline. We plot the raw change-from-baseline scores used to compute Cohen’s *d* metrics for (a) learned ABC-mSW total, (b) standard ABC-mSW total, (c) learned SRS2-ER total, and (d) standard SRS2-ER total. Weights of all items were normalized to sum to one for direct comparison between learned and standard scores. The learned scores diverged during the randomized phases, consistent with significant mean Cohen’s *d* values. The placebo group then converged to the original oxytocin group once all participants received oxytocin in the open-label phase. By contrast, the standard scores exhibited no significant group differences in the randomized phase. Lines denote group means, and error bars indicate one standard error of the mean. All panels share the same legend.

### Other Open-Label Extension Results

The original ABC-mSW and SRS2-ER total scores both declined significantly in the open-label phase (ABC-mSW: mean Cohen’s *d* = −0.31, *p* = 0.009); SRS2-ER: mean Cohen’s *d* = −0.29, *p* = 0.002). None of these effects replicated in the randomized phase.

**Supplementary Fig. 2:**
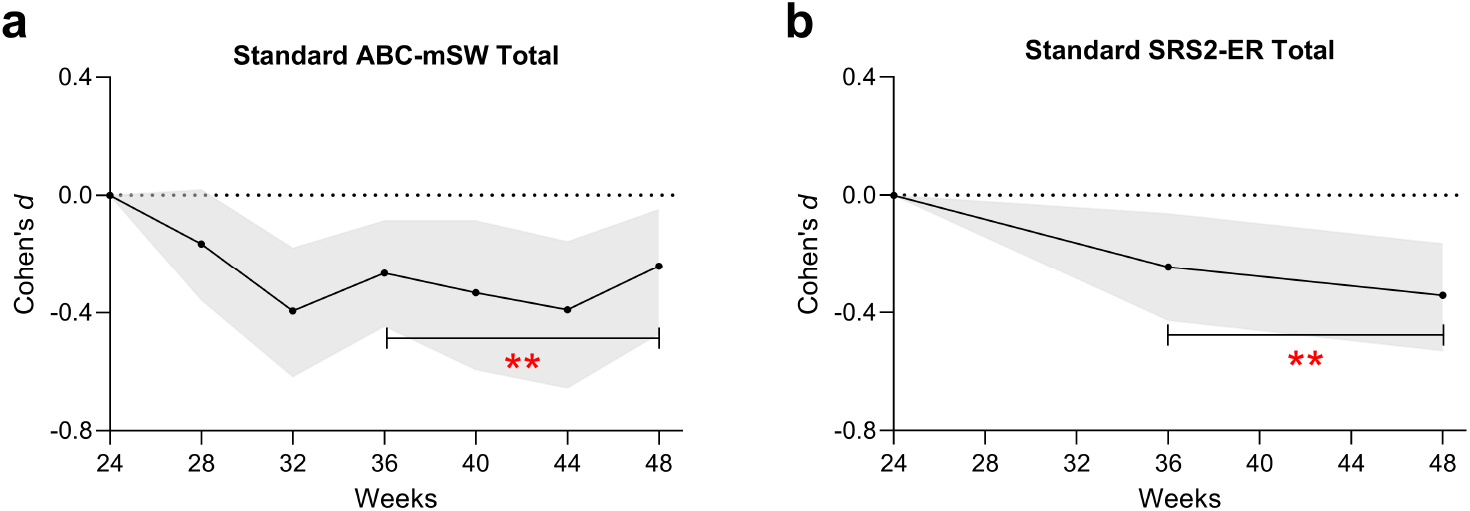
Effects in the open-label extension phase. (a) Patients who cross-over from placebo to oxytocin exhibited reduced maladaptive behaviors with the standard ABC-mSW total score at weeks 36-48. (b) Similarly, oxytocin was associated with significant improvements in both maladaptive and adaptive behaviors based on the standard SRS2-ER total score. However, these results did not hold in the randomized phase.

